# Cumulative Incidence of Motor and Cognitive Features in the ALS-FTD Spectrum

**DOI:** 10.1101/2024.04.30.24306638

**Authors:** Barbara E. Spencer, Sharon X. Xie, Daniel T. Ohm, Lauren Elman, Colin C. Quinn, Defne Amado, Michael Baer, Edward B. Lee, Vivianna M. Van Deerlin, Laynie Dratch, Lauren Massimo, David J. Irwin, Corey T. McMillan

**Author notes:** Correspondence to: Corey T. McMillan Richards Laboratories 3700 Hamilton Walk Suite 601 B Philadelphia, PA, 19104.

## Abstract

In frontotemporal degeneration (FTD) and amyotrophic lateral sclerosis (ALS), subsequent motor or cognitive-behavioral features, respectively, are associated with shorter survival. However, factors influencing subsequent feature development remain largely unexplored. In this study, we examined whether the presence of a *C9orf72* expansion or the initial clinical syndrome was associated with increased risk of subsequent feature development in individuals with ALS and FTD.

We performed a retrospective evaluation of the entire disease course of individuals with ALS and FTD who had neuropathological confirmation of TDP-43 proteinopathy at autopsy or a *C9orf72* hexanucleotide repeat expansion. We examined the odds and hazard of subsequent feature development and assessed whether each was modified by the presence of a *C9orf72* expansion or initial clinical syndrome. At autopsy, we evaluated the association between TDP-43 pathology burden in characteristic brain regions and features across the FTD-ALS spectrum.

For individuals with ALS (n=168) and FTD (n=73), binary logistic regression revealed increased odds (OR=3.49[95% CI 1.64-7.80], p=0.002) for developing subsequent features in those with a *C9orf72* expansion compared to those without and decreased odds (OR=0.25[95% CI 0.12-0.53], p<0.001) for developing subsequent features in those with an initial ALS clinical syndrome compared to those with an initial FTD clinical syndrome. Cox proportional hazard analyses revealed an increased hazard (HR=3.78[95% CI 1.86-7.65], p<0.001) for developing subsequent features in those with a *C9orf72* expansion compared to those without. We observed a 94-month difference in the time after symptom onset of the initial clinical syndrome that a given person without a *C9orf72* expansion reached the highest probability of developing subsequent features (0.12[95% CI (0.03-0.19], 113.00 months) and a person with a *C9orf72* expansion surpassed that probability (0.13[95% CI 0.06-0.19], 19.00 months). Beyond *C9orf72* expansion status, cox proportional hazard analyses revealed a decreased hazard (HR=0.48[95% CI 0.25-0.95], p=0.03) for developing subsequent features in those with an initial ALS clinical syndrome compared to those with an initial FTD clinical syndrome. Age at symptom onset and sex were not associated with development of subsequent features. The distribution of TDP-43 pathology across characteristic brain regions reflected both the initial clinical syndrome and subsequent features, with relatively preserved spinal cord only in FTD cases without subsequent motor features (p<0.0001) and relatively preserved neocortical regions only in ALS cases without subsequent cognitive-behavioral features (p<0.0001).

These data highlight the need for clinician vigilance to detect the onset of subsequent motor and cognitive-behavioral features in patients carrying a *C9orf72* expansion, regardless of initial clinical syndrome. *C9orf72* clinical care can be enhanced through coordination between cognitive and neuromuscular clinics.

**Abbreviated Summary:** Spencer et al. demonstrated both the presence of a C9orf72 expansion and the initial clinical syndrome modify risk of subsequent feature development in frontotemporal degeneration and amyotrophic lateral sclerosis, highlighting the need for clinician vigilance to detect the onset of subsequent motor and cognitive-behavioral features in this disease spectrum.

## Introduction

Frontotemporal degeneration (FTD) and amyotrophic lateral sclerosis (ALS) are multi-system disorders that occur along a spectrum of cognitive-behavioral and neuromuscular impairments, respectively.^1^ Pathologic, genetic, and clinical features support the existence of this clinicopathologic spectrum. First, TAR DNA-binding protein ∼43kDa (TDP-43) inclusions are the pathological hallmark of approximately 50% of FTD and the vast majority ( >98%) of ALS cases.^2^ Second, hexanucleotide repeat expansions in *C9orf72*^3,4^ can cause FTD, ALS, or both, and account for ∼5% of simplex and one-third of familial FTD and ALS cases.^5,6^ Third, clinical overlap between FTD and ALS can occur, even in the absence of a *C9orf72* expansion, though estimates of the relative frequency of subsequent feature development vary widely.

In both FTD and ALS, the presence of subsequent motor or cognitive-behavioral features, respectively, is associated with shorter survival.^7–10^ Therefore, understanding the risk of subsequent features within FTD and ALS is critical, yet factors influencing the risk of subsequent feature development remain largely unexplored.

Previous studies estimate that ∼9-15% of individuals with ALS eventually develop frank FTD, though 30-50% develop some level of cognitive impairment.^1,11–15^ Recently, using appropriate normative data, we found that ∼15% of individuals with ALS have cognitive impairment.^16^ Likewise, ∼4-14% of individuals with FTD develop a subsequent motor impairment consistent with ALS,^17–20^ a number that may underestimate the incidence since latent evidence of neuromuscular impairment is rarely assessed. Estimates of subsequent motor or cognitive-behavioral feature development specifically in those with disease due to a *C9orf72* expansion vary widely (∼11-60%).^3,21–25^ It is difficult to compare estimates across studies which use different criteria to determine the presence of subsequent features. In several studies that directly compare persons with *C9orf72* expansions to those without *C9orf72* expansions, a higher frequency of subsequent feature development has been reported in individuals with *C9orf72* expansions (23-50%) compared to those without *C9orf72* expansions (4-12%). ^26–28^ However, research limited to individuals with *C9orf72* expansions likely overestimate concomitant FTD and ALS.

A further major limitation of prior research evaluating subsequent feature development in FTD and ALS is the reliance on clinically-rather than neuropathologically-defined cases. Upper and/or lower motor neuron features are extraordinarily rare in FTD due to underlying tau pathology.^29,30^ Tau pathology accounts for nearly half of all FTD cases and is not easily clinically differentiated from FTD due to underlying TDP-43. Thus, subsequent ALS in FTD is likely underestimated when individuals without an underlying TDP-43 proteinopathy are included in risk models.

Recent evidence suggests that the risk of developing subsequent ALS in individuals with an initial presentation of FTD decreases with time from FTD symptom onset.^27^ However, we are unaware of studies evaluating the hazard of subsequent feature development over the disease course, which is an important consideration for patient prognostication. Further, the hazard of subsequent feature development stratified by initial clinical syndrome, FTD or ALS, has not yet been explored.

In this study, we examine whether the presence of a *C9orf72* expansion is associated with increased risk of subsequent feature development in individuals with ALS, excluding those with *SOD1* related disease lacking TDP-43 pathology, and FTD, excluding those with non-TDP-43 proteinopathy at autopsy. We further ask whether the risk of subsequent feature development is modified by the initial clinical syndrome, FTD or ALS, beyond *C9orf72* expansion status. Using a targeted, retrospective evaluation of patients with a confirmed TDP-43 proteinopathy who were followed clinically from initial diagnosis until death, we are able to assess the presence or absence of subsequent feature development through the entire disease course. At autopsy, we evaluate the distribution of TDP-43 pathology in characteristic brain regions to evaluate the association between pathology and features across the FTD-ALS spectrum.

## Materials and methods

### Participants

We retrospectively evaluated data from 241 deceased individuals from the University of Pennsylvania Integrated Neurodegenerative Disease Database.^31,32^ Informed consent was obtained for all participants through a procedure approved by an Institutional Review Board convened at the University of Pennsylvania.

Inclusion was limited to individuals with:

1. An initial clinical syndrome consistent with ALS or FTD as evaluated by a board-certified neurologist with expertise in neuromuscular and/or cognitive disorders, and
2. Complete records for date of birth, date of symptom onset for the initial clinical syndrome, and date of death, and
3. Blood DNA or postmortem frozen brain tissue screened for presence or absence of a *C9orf72* expansion (see *Genetic analysis*), and
4. Neuropathological confirmation of a TDP-43 proteinopathy at autopsy (see *Neuropathological evaluation*) *OR* the presence of a *C9orf72* expansion, and
5. Chart review

Two qualified reviewers performed a retrospective chart review of all available electronic and paper health records for each individual. On average individuals with ALS were seen clinically every 3 months and those with FTD were seen clinically every 6 months. Each reviewer independently assessed each case and recorded the presence or absence of subsequent features during disease course and, if present, the date of onset of subsequent features. Discrepancies between reviewers were reconciled by a third reviewer, and Cohen’s Kappa was used to measure inter-rater reliability. Subsequent features were defined as any clinical evidence of motor impairment in an individual with an initial clinical presentation of FTD or any clinical evidence of cognitive-behavioral impairment in an individual with an initial clinical presentation of ALS. Subsequent motor features included irregular or slowed gait, foot drop, dysphagia, dysarthria, fasciculations, limb weakness, and shortness of breath. Subsequent cognitive-behavioral features included personality change, disinhibition, apathy, compulsive or ritualistic behaviors, agitation, deficits in executive function, lack of empathy, and lack of insight. We did not consider the following as subsequent features: Parkinsonism such as rigidity, pseudobulbar features like inappropriate laughter or crying, or decades long behavioral features lacking clear evidence of progression. When month was not determinable the default of January was used, and when day was not determinable the first of the month was used. Time to subsequent feature development was calculated as the difference in months between the date of symptom onset of the initial clinical syndrome and the date of onset of subsequent features.

### Genetic analysis

DNA was extracted from peripheral blood or frozen brain tissue following the manufacturer’s protocols (QuickGene DNA whole blood kit (Autogen) for blood, and QIAamp DNA Mini Kit (Qiagen) for brain tissue). All individuals were tested for *C9orf72* hexanucleotide repeat expansions using a modified repeat-primed PCR as previously described. Cases were defined as having a pathogenic *C9orf72* expansion if >30 hexanucleotide repeats were identified. Individuals were further screened for pathogenic variants associated with FTD and/or ALS using exome/genome sequencing, and/or a custom targeted multi neurodegenerative disease sequencing panel.^31^ The sequencing data was analyzed using Geneticist Assistant software (Soft Genetics, State College, PA).

### Neuropathological evaluation

Detailed neuropathological assessments were performed using established and uniform methods of fixation, tissue processing, IHC with well-characterized antibodies, and current neuropathological criteria, as has been described in detail elsewhere.^31,33,34^ Briefly, 15 brain regions from one hemisphere, alternating right and left at random, are routinely sampled at autopsy, formalin-fixed, and processed for immunohistochemical staining using 1D3 (gift of Manuela Neumann and Elisabeth Kremmer) for phosphorylated TDP-43. Each brain region was semi-quantitatively scored for the burden of TDP-43 (0, absent; 0.5, rare; 1, mild; 2, moderate; 3, severe, Supplementary Figure 1).

### Statistical analysis

We first assessed the effect of *C9orf72* expansion status (present or absent) and initial clinical syndrome (FTD or ALS) on the odds of developing subsequent features at any point from symptom onset of the initial clinical syndrome until death using binary logistic regression, controlling for age at initial clinical syndrome onset and sex. We next assessed the effect of *C9orf72* expansion status, alone and in combination with initial clinical syndrome on subsequent feature development using a time-to-event model.

The Kaplan-Meier method was used to estimate the cumulative incidence of subsequent feature development. An individual was considered at risk for subsequent feature development from the time of symptom onset of the initial clinical syndrome until death. We subsequently considered an individual at risk until either death, tracheostomy, or permanent assisted ventilation, consistent with the definition of survival in some clinical trials.^35^ Here, permanent assisted ventilation was defined as the date at which the individual reported using noninvasive positive pressure ventilation for more than 23 hours per day.

Cox proportional hazard analyses, controlling for age at initial clinical syndrome onset and sex, were used to examine the influence of *C9orf72* expansion status, alone and in combination with initial clinical syndrome, on subsequent feature development. The proportional hazards assumption was tested for each Cox regression model fit.^36^

Finally, the distribution of TDP-43 pathology was evaluated across the FTD-ALS spectrum, using FDR-adjusted Kruskal-Wallis tests to evaluate the difference in TDP-43 burden in each characteristic brain region across individuals grouped by initial clinical syndrome and subsequent features, followed by FDR-adjusted pairwise Dunn’s tests to identify which groups were different. All statistical tests were two-sided. All analysis was done using R (version 4.2.3).

### Data availability

Diagnosis, genetic, and neuropathological data may be requested and upon approval of reasonable requests may be shared with individual investigators. Data requests can be completed through the Penn Neurodegenerative Data Sharing Committee webform: https://www.pennbindlab.com/data-sharing

## Results

### Subsequent feature development

73 individuals with initial FTD (35 [48%] *C9orf72* expansion present) and 168 individuals with initial ALS (65 [39%] *C9orf72* expansion present) were included in the study. Demographic, clinical, and genetic characteristics for all individuals are summarized in Table 1. Of the 73 individuals with initial FTD, 22 (30%) developed subsequent motor features. Of the 168 individuals with initial ALS, 16 (10%) developed subsequent cognitive-behavioral features. Cohen’s Kappa indicated very high agreement in the assessment of each case for the presence or absence of subsequent features (0.84). Of the 38 individuals who developed subsequent features, regardless of *C9orf72* status, 27 did so prior to the discovery of *C9orf72* expansions in FTD and ALS in 2011.^3,4^ Of the 26 individuals with a *C9orf72* expansion who developed subsequent features, 16 did so prior to 2011.

**Table 1.**
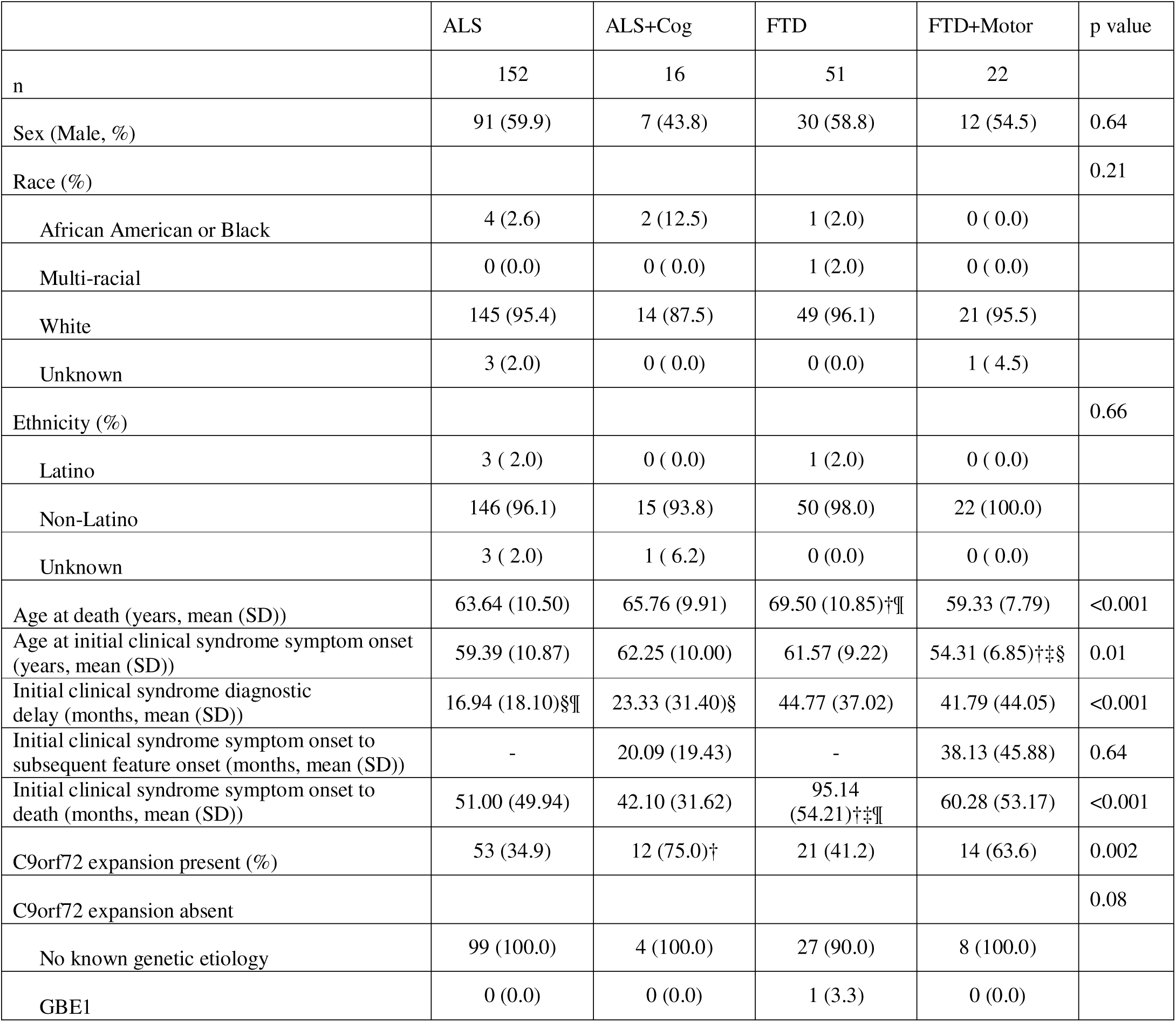
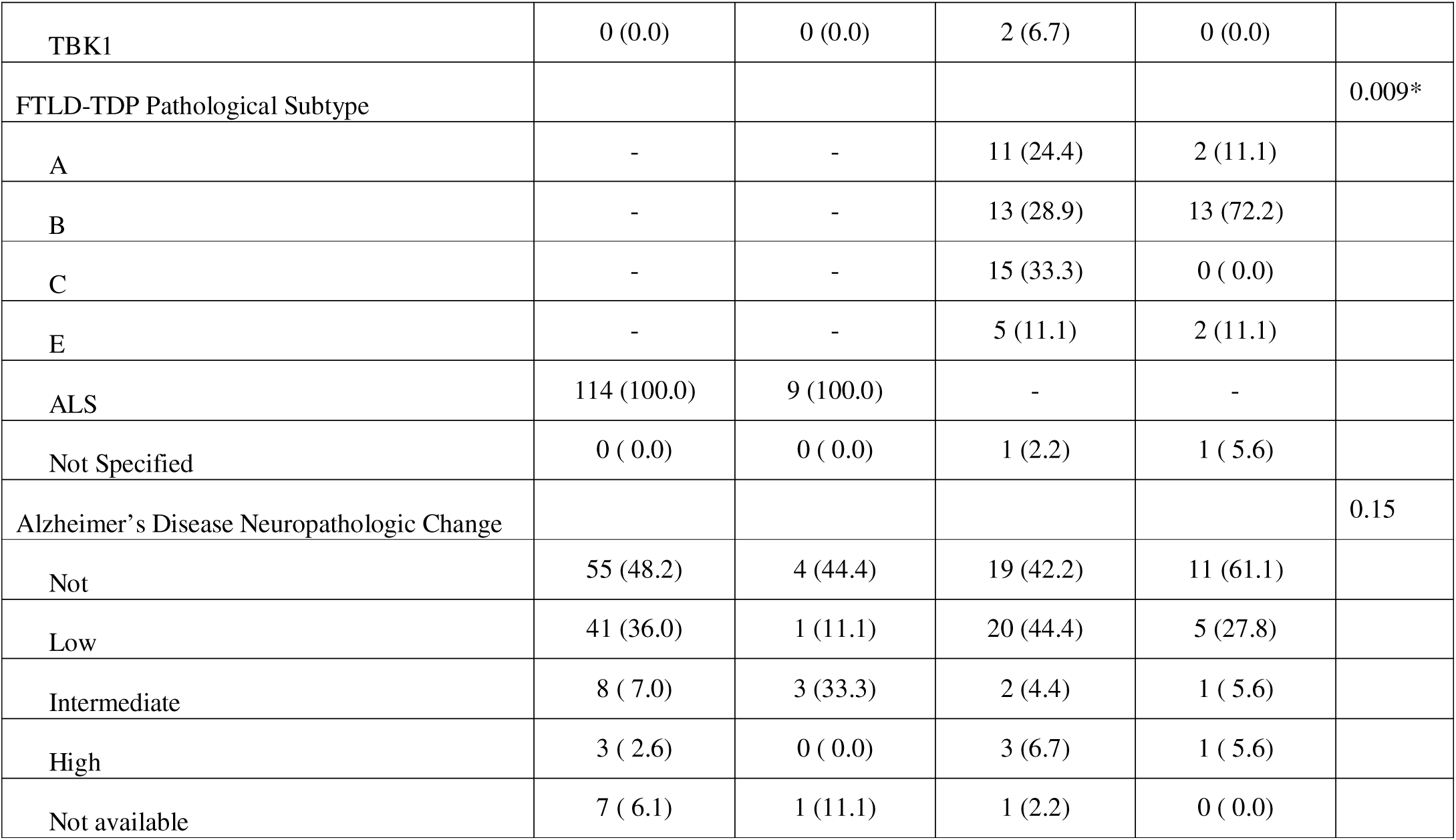
Sample Characteristics. Demographic, clinical, and genetic data for all individuals, split by initial clinical syndrome (FTD or ALS) and the presence of subsequent motor (FTD+Motor) or cognitive-behavioral (ALS+Cog) features. For continuous variables, mean and standard deviation are provided; p values represent Kruskal-Wallis tests for group comparisons, and differences from †ALS, ‡ALS+Cog, §FTD, or ¶FTD+Motor based on FDR-adjusted significant pairwise Dunn’s tests are indicated. For categorical variables, count and percentage are provided; p values represent χ^2^ tests for group comparisons, and differences from †ALS, ‡ALS+Cog, §FTD, or ¶FTD+Motor based on FDR-adjusted significant pairwise χ^2^ tests are indicated. *comparison between FTD and FTD+Motor only

|  | ALS | ALS+Cog | FTD | FTD+Motor | p value |
| --- | --- | --- | --- | --- | --- |
| n | 152 | 16 | 51 | 22 |  |
| Sex (Male, %) | 91 (59.9) | 7 (43.8) | 30 (58.8) | 12 (54.5) | 0.64 |
| Race (%) |  |  |  |  | 0.21 |
| African American or Black | 4 (2.6) | 2 (12.5) | 1 (2.0) | 0 (0.0) |  |
| Multi-racial | 0 (0.0) | 0 (0.0) | 1 (2.0) | 0 (0.0) |  |
| White | 145 (95.4) | 14 (87.5) | 49 (96.1) | 21 (95.5) |  |
| Unknown | 3 (2.0) | 0 (0.0) | 0 (0.0) | 1 (4.5) |  |
| Ethnicity (%) |  |  |  |  | 0.66 |
| Latino | 3 (2.0) | 0 (0.0) | 1 (2.0) | 0 (0.0) |  |
| Non-Latino | 146 (96.1) | 15 (93.8) | 50 (98.0) | 22 (100.0) |  |
| Unknown | 3 (2.0) | 1 (6.2) | 0 (0.0) | 0 (0.0) |  |
| Age at death (years, mean (SD)) | 63.64 (10.50) | 65.76 (9.91) | 69.50 (10.85)†¶ | 59.33 (7.79) | <0.001 |
| Age at initial clinical syndrome symptom onset (years, mean (SD)) | 59.39 (10.87) | 62.25 (10.00) | 61.57 (9.22) | 54.31 (6.85)†‡§ | 0.01 |
| Initial clinical syndrome diagnostic delay (months, mean (SD)) | 16.94 (18.10)§¶ | 23.33 (31.40)§ | 44.77 (37.02) | 41.79 (44.05) | <0.001 |
| Initial clinical syndrome symptom onset to subsequent feature onset (months, mean (SD)) | - | 20.09 (19.43) | - | 38.13 (45.88) | 0.64 |
| Initial clinical syndrome symptom onset to death (months, mean (SD)) | 51.00 (49.94) | 42.10 (31.62) | 95.14 (54.21)†‡¶ | 60.28 (53.17) | <0.001 |
| C9orf72 expansion present (%) | 53 (34.9) | 12 (75.0)† | 21 (41.2) | 14 (63.6) | 0.002 |
| C9orf72 expansion absent |  |  |  |  | 0.08 |
| No known genetic etiology | 99 (100.0) | 4 (100.0) | 27 (90.0) | 8 (100.0) |  |
| GBE1 | 0 (0.0) | 0 (0.0) | 1 (3.3) | 0 (0.0) |  |
| FTD-ALS subsequent feature development |  |  |  |  |  |
| TBK1 | 0 (0.0) | 0 (0.0) | 2 (6.7) | 0 (0.0) |  |
| FTLD-TDP Pathological Subtype |  |  |  |  | 0.009* |
| A | - | - | 11 (24.4) | 2 (11.1) |  |
| B | - | - | 13 (28.9) | 13 (72.2) |  |
| C | - | - | 15 (33.3) | 0 ( 0.0) |  |
| E | - | - | 5 (11.1) | 2 (11.1) |  |
| ALS | 114 (100.0) | 9 (100.0) | - | - |  |
| Not Specified | 0 ( 0.0) | 0 ( 0.0) | 1 (2.2) | 1 ( 5.6) |  |
| Alzheimer's Disease Neuropathologic Change |  |  |  |  | 0.15 |
| Not | 55 (48.2) | 4 (44.4) | 19 (42.2) | 11 (61.1) |  |
| Low | 41 (36.0) | 1 (11.1) | 20 (44.4) | 5 (27.8) |  |
| Intermediate | 8 ( 7.0) | 3 (33.3) | 2 (4.4) | 1 ( 5.6) |  |
| High | 3 ( 2.6) | 0 ( 0.0) | 3 (6.7) | 1 ( 5.6) |  |
| Not available | 7 ( 6.1) | 1 (11.1) | 1 (2.2) | 0 ( 0.0) |  |

#### Logistic regression

Binary logistic regression revealed increased odds of developing subsequent features in those with *C9orf72* expansions relative to those without (OR= 3.49 [95% CI 1.64-7.80], p=0.002) and decreased odds of developing subsequent features in those with an initial ALS clinical syndrome compared to those with an initial FTD clinical syndrome (OR= 0.25 [95% CI 0.12-0.53], p<0.001). Neither age at initial clinical syndrome onset (OR=0.98 [95% CI 0.95-1.02], p=0.45) nor sex (OR=0.65 [95% CI 0.31-1.38], p=0.26) contributed to the odds of developing subsequent features.

#### Cox proportional hazards

Those with *C9orf72* expansions had an increased risk for developing subsequent features compared to those without (HR= 3.78 [95% CI 1.86-7.65], p<0.001). Kaplan-Meier cumulative incidence plots, stratified by *C9orf72* expansion status alone or in combination with with initial clinical syndrome are displayed in Figure 1. In persons with *C9orf72* expansions, the probability of a given person developing subsequent features reached 0.5 (50%) at 168 months (14 years) after symptom onset of the initial clinical syndrome (Figure 1). In persons without *C9orf72* expansions, the time at which the probability of a given person developing subsequent features would reach 0.5 is unable to be estimated due to the infrequent development of subsequent features in this group. In combination with *C9orf72* carrier status, an initial ALS clinical syndrome was associated with a decreased hazard for developing subsequent features (HR= 0.48 [95% CI 0.25-0.95], p=0.03). Neither age at initial clinical syndrome onset (HR range across models 0.99-1.00 [95% CI range across models 0.96-1.03]) nor sex (HR range across models 0.74-0.75 [95% CI range across models 0.39-1.43]) were associated with the hazard for developing subsequent features in either model. Tracheostomy was administered to 6 of 241 participants (2.49%) and 6 of 241 participants (2.49%) used permanent assisted ventilation. When we defined survival by death, tracheostomy, or permanent assisted ventilation, we observed similar results to those using death only; individuals with *C9orf72* expansions had an increased risk for developing subsequent features compared to those without (HR= 3.77 [95% CI 1.86-7.65], p<0.001), and, in combination with *C9orf72* carrier status, an initial ALS clinical syndrome was associated with a decreased hazard for developing subsequent features (HR= 0.49 [95% CI 0.25–0.97], p=0.04).

**Figure 1.**
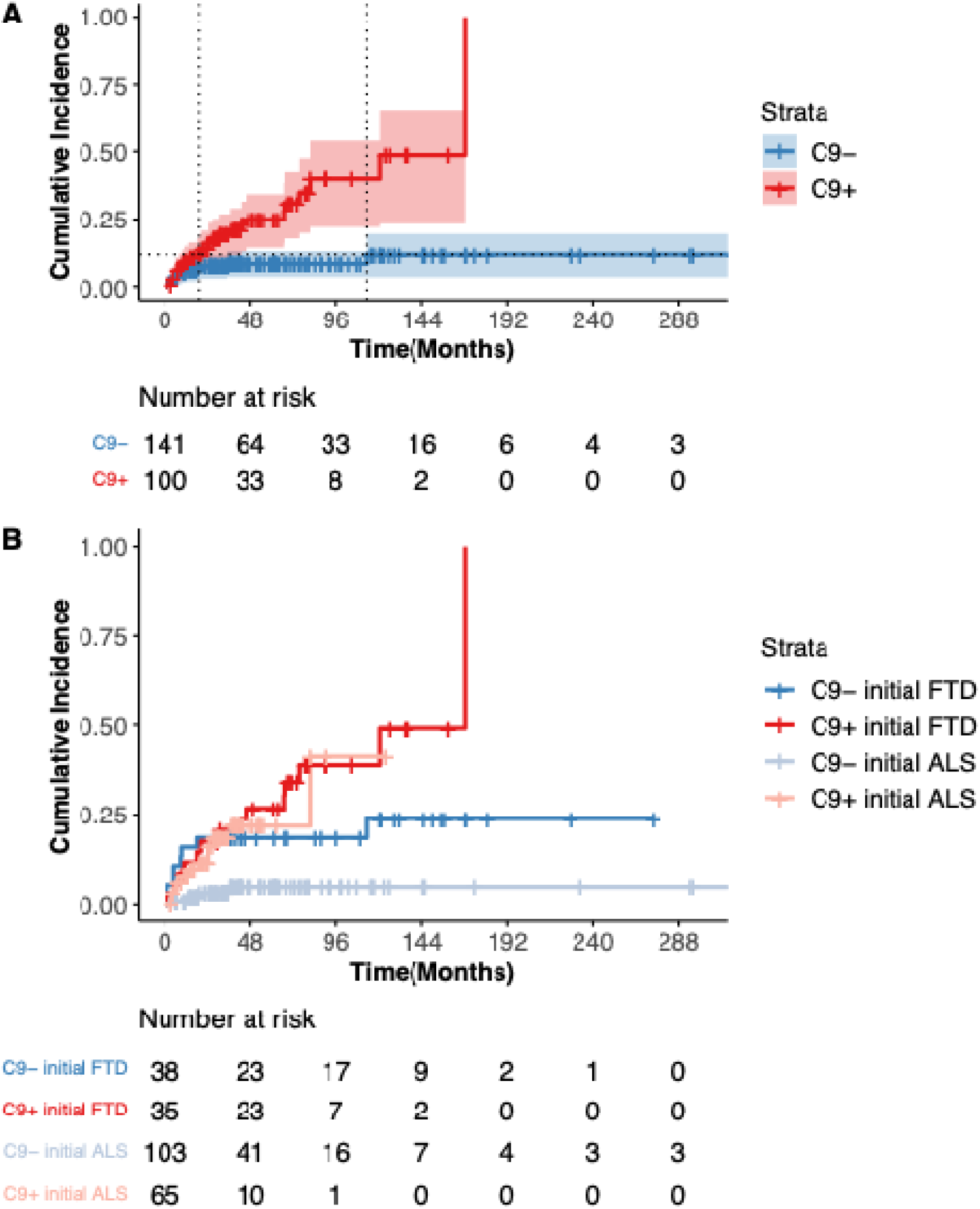
C9orf72 hexanucleotide repeat expansions increase risk for subsequent feature development. Kaplan-Meier cumulative incidence plots are displayed stratified by (**A**) the absence (blue, N=141) or presence (red, N=100) of a *C9orf72* expansion alone and (**B**) combined with initial clinical syndrome: initial clinical syndrome of FTD without *C9orf72* expansion (blue, N=38), initial clinical syndrome of FTD with *C9orf72* expansion (red, N=35), initial clinical syndrome of ALS without *C9orf72* expansion (light blue, N=103), and initial clinical syndrome of ALS with *C9orf72* expansion (salmon, N=65). Vertical rises indicate the development of subsequent cognitive-behavioral or motor features. Tick marks indicate censoring due to death. Shading represents 95% confidence intervals. The table shows the number of living individuals in the sample who have not yet developed subsequent features at a given time point. In (A) dotted horizontal line represents the highest probability of developing subsequent features reached for a given person without a *C9orf72* expansion in our sample (cumulative incidence 0.12); dotted vertical lines represent the time after symptom onset of the initial clinical syndrome a given person with (19 months) or without (113 months) a *C9orf72* expansion reaches this probability.

### Regional TDP-43 burden

At autopsy, the distribution of TDP-43 pathology across characteristic brain regions reflected both the initial clinical syndrome and subsequent features. First, this was done in characteristic FTD regions, specifically the cingulate gyrus, middle frontal cortex, angular gyrus, and superior/middle temporal cortex (Figure 2). Pairwise Dunn’s tests revealed a lower burden of TDP-43 pathology in ALS cases without subsequent cognitive-behavioral features compared to FTD cases (with or without subsequent motor features) across all characteristic FTD regions (all FDR-adjusted p<.001). Further, a lower burden of TDP-43 pathology was observed in ALS cases without subsequent cognitive-behavioral features compared to ALS cases with subsequent cognitive-behavioral features across all characteristic FTD regions (all FDR-adjusted p<.05). Second, we examined TDP-43 burden in characteristic ALS regions, motor cortex and spinal cord. In the spinal cord, a lower burden of TDP-43 pathology was observed in FTD cases without subsequent motor features compared to ALS cases (with or without subsequent cognitive-behavioral features) and FTD cases with subsequent motor features (all FDR-adjusted p<.01). However, no significant differences in TDP-43 burden were observed in the motor cortex across individuals grouped by initial clinical syndrome and subsequent features using Kruskal-Wallis test (p=.10).

**Figure 2.**
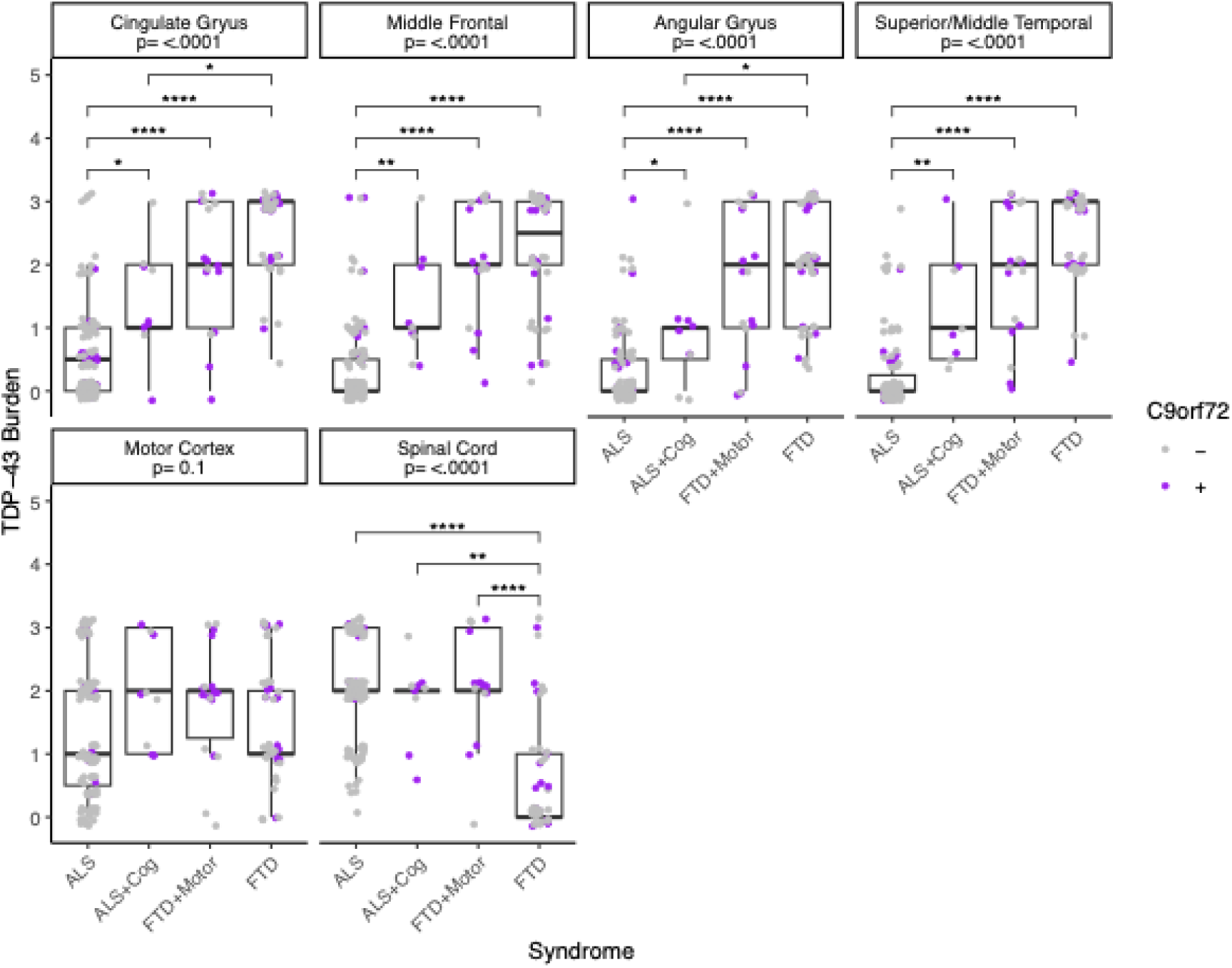
Regional TDP-43 burden reflects initial clinical syndrome and subsequent features. The burden of TDP-43 pathology was examined across individuals grouped by initial clinical syndrome and subsequent features. This was done in characteristic FTD regions (top row: cingulate gyrus (N=175), middle frontal cortex (N=178), angular gyrus (N=175), and superior/middle temporal cortex (N=177)), and characteristic ALS regions (bottom row: motor cortex (N=179) and spinal cord (N=176)). The FDR adjusted p-values for each Kruskal-Wallis test is displayed by region. Asterisks represent FDR-adjusted significant pairwise Dunn’s tests within region (****p<.0001, ***p<.001, **p<.01, *p<.05). Jitter was added around the semi-quantitatively scored TDP-43 burden (0, absent; 0.5, rare; 1, mild; 2, moderate; 3, severe) for individual data point visualization. Individual data points were colored by the absence (gray) or presence (purple) of a *C9orf72* expansion.

## Discussion

Individuals with an initial clinical ALS or FTD syndrome with an underlying TDP-43 proteinopathy are at risk for developing subsequent cognitive-behavioral or motor features, respectively, and this risk is heightened for those with disease due to a *C9orf72* hexanucleotide repeat expansion. Regardless of genetic status, there is a modest risk for the development of subsequent features in the first year after symptom onset of the initial clinical syndrome, with 9.0% of persons with *C9orf72* expansions and 5.7*%* of those without *C9orf72* expansions developing subsequent features in this timeframe. However, by death, 26.0% of those with *C9orf72* expansions develop subsequent features compared to only 8.5*%* of those without *C9orf72* expansions, in line with previous estimates.^26–28^ While some studies report a median interval of less than one year between the onset of the initial clinical syndrome and subsequent features in those with a *C9orf72* expansion, we observed a median of just under 2 years (22 months).^37^ In our sample, a given person without a *C9orf72* expansion reaches a probability of developing subsequent features of 12% at 113 months after symptom onset of the initial clinical syndrome, after which there is no increase in probability. In contrast, for a given person with a *C9orf72* expansion, that probability is surpassed by 19 months, a 94-month difference. By 168 months (14 years) after symptom onset of the initial clinical syndrome, a person with a *C9orf72* expansion has a 50% probability of developing subsequent features. The mean time between symptom onset of the initial clinical syndrome and death is shorter than 168 months across ALS and FTD groups, and few individuals remain at risk for subsequent feature development at this time point. However, we observed two persons with *C9orf72* expansions who develop subsequent features 10+ years after symptom onset of the initial clinical syndrome, similar to some previously reported intervals.^38^

In this study, we found that the cumulative incidence of subsequent feature development was lower for individuals with an initial ALS clinical syndrome, beyond the effect of *C9orf72* expansion. The time between symptom onset of the initial clinical syndrome and death was shortest for individuals with initial ALS, with or without subsequent feature development, aligning with prior work that found that long-term survivors in the FTD-ALS spectrum were more likely to be individuals with cognitive symptoms at onset.^39^ This difference in survival may partially explain the lower cognitive-behavioral feature development we observed in ALS, although there was no statistical difference in survival between ALS and FTD with subsequent motor features. It is also possible that it is easier for cognitive neurologists to reliably observe motor features in the context of FTD than it is for neuromuscular neurologists to detect cognitive-behavioral features in the context of ALS.

The presence of subsequent feature development was associated with a shorter interval between symptom onset of the initial clinical syndrome and death for both initial FTD and ALS, aligning with previous research,^7–10^ although only individuals with initial FTD without subsequent motor features lived significantly longer than other groups in our sample. We also observed a younger age at onset of cognitive-behavioral symptoms for individuals with initial FTD who develop subsequent motor features (54.31 [6.85]) than those who never develop subsequent features (61.57 [9.22], Table 1).

At autopsy, the distribution of TDP-43 pathology in characteristic regions reflected both the initial syndrome and subsequent features, with relatively preserved spinal cord only in FTD cases without subsequent motor features and relatively preserved neocortical regions only in ALS cases without subsequent cognitive-behavioral features. These data align with previous work highlighting the distribution and severity of TDP-43 pathology relates to the clinical expression of dementia and motor impairments across the spectrum of TDP-43 proteinopathies^40–42^ and provides important cross-validation for our clinical designations of ALS, FTD, and subsequent features. As expected, we did not observe any occurrences of FTD with subsequent motor features in FTLD-TDP type C.

Future studies are necessary to determine a biological explanation for why individuals with a *C9orf72* expansion have a higher risk for developing subsequent features relative to those without despite sharing common underlying TDP-43 pathology. More people without *C9orf72* expansions than those with them may ultimately develop subsequent features, despite subsequent feature development being a lower probability event, as most individuals with ALS and FTD do not have a *C9orf72* expansion, and studies are needed to elucidate risk factors in this population.^43–45^

As with all epidemiological studies there are several caveats to consider in our approach. In order to ensure TDP-43 pathology in this study, we relied on neuropathological confirmation, which necessitated a retrospective evaluation of cases. One should consider any potential sources of bias introduced by our primary focus on individuals who participated in brain donation, who tend to be more educated and non-representative of the population per se. Prospective collection of both reported feature onset as well as detailed electrophysiological support and neuropsychological testing would strengthen the study; however, our postmortem data support our clinical groupings (Figure 2). It is also necessary for future studies to address other potential modifiers of risk for concomitant cognitive-behavioral and motor impairment such as cognitive and motor reserve factors^46^ and other environmental^47^ and genetic^48,49^ modifiers.

These findings should encourage clinicians to have elevated vigilance for the onset of subsequent cognitive-behavioral and motor features in patients with a *C9orf72* expansion regardless of initial clinical syndrome and may warrant dual referrals between cognitive and neuromuscular clinics. Early work likely failed to observe an association between subsequent features and survival in ALS due to a focus on memory rather than FTD-like cognitive/behavioral impairment.^12^ Therefore, it is critical to properly evaluate these patients. This information may also be helpful for patients and families in terms of prognosis and understanding the implications of a *C9orf72* expansion in their family.

## Acknowledgements

The authors would like to thank Laura Hennessy, Katerina Placek, Carrie Caswell, and Kaylee Naczi who contributed to the recruitment, data collection, and characterization of individuals included in this study.

## Funding

National Institutes of Health grant F32AG079618 (BES), P01AG066597 (CTM), NS109260 (DJI), P30AG072979 (EBL), Penn Institute on Aging, and The DeCrane Family Fund for Primary Progressive Aphasia

## Competing interests

EBL reports consulting fees from WaveBreak Therapeutics. LD reports consulting fees from Passage Bio, Biogen, and Sano Genetics. The authors report no other relevant competing interests.

## Notes

### Funding Statement

National Institutes of Health grant F32AG079618 (BES), P01AG066597 (CTM), NS109260 (DJI), Penn Institute on Aging, and The DeCrane Family Fund for Primary Progressive Aphasia

### Summary of Updates

This version of the manuscript has been revised.

## References

1. Strong MJ, Abrahams S, Goldstein LH, et al. Amyotrophic lateral sclerosis - frontotemporal spectrum disorder (ALS-FTSD): Revised diagnostic criteria. Amyotrophic Lateral Sclerosis and Frontotemporal Degeneration. 2017;18(3-4):153–174. doi:10.1080/21678421.2016.1267768

2. Neumann M, Sampathu DM, Kwong LK, et al. Ubiquitinated TDP-43 in Frontotemporal Lobar Degeneration and Amyotrophic Lateral Sclerosis. Science. 2006;314(5796):130–133. doi:10.1126/science.1134108

3. DeJesus-Hernandez M, Mackenzie IR, Boeve BF, et al. Expanded GGGGCC hexanucleotide repeat in non-coding region of C9ORF72 causes chromosome 9p-linked frontotemporal dementia and amyotrophic lateral sclerosis. Neuron. 2011;72(2):245–256. doi:10.1016/j.neuron.2011.09.011

4. Renton AE, Majounie E, Waite A, et al. A hexanucleotide repeat expansion in C9ORF72 is the cause of chromosome 9p21-linked ALS-FTD. Neuron. 2011;72(2):257–268. doi:10.1016/j.neuron.2011.09.010

5. Majounie E, Renton AE, Mok K, et al. Frequency of the C9orf72 hexanucleotide repeat expansion in patients with amyotrophic lateral sclerosis and frontotemporal dementia: a cross-sectional study. Lancet Neurol. 2012;11(4):323–330. doi:10.1016/S1474-4422(12)70043-1

6. Zou ZY, Zhou ZR, Che CH, Liu CY, He RL, Huang HP. Genetic epidemiology of amyotrophic lateral sclerosis: a systematic review and meta-analysis. J Neurol Neurosurg Psychiatry. 2017;88(7):540–549. doi:10.1136/jnnp-2016-315018

7. Olney RK, Murphy J, Forshew D, et al. The effects of executive and behavioral dysfunction on the course of ALS. Neurology. 2005;65(11):1774–1777. doi:10.1212/01.wnl.0000188759.87240.8b

8. Josephs KA, Knopman DS, Whitwell JL, et al. Survival in two variants of tau-negative frontotemporal lobar degeneration: FTLD-U vs FTLD-MND. Neurology. 2005;65(4):645–647. doi:10.1212/01.wnl.0000173178.67986.7f

9. Hu WT, Shelnutt M, Wilson A, et al. Behavior matters--cognitive predictors of survival in amyotrophic lateral sclerosis. PLoS One. 2013;8(2):e57584. doi:10.1371/journal.pone.0057584

10. Hodges JR, Davies R, Xuereb J, Kril J, Halliday G. Survival in frontotemporal dementia. Neurology. 2003;61(3):349–354. doi:10.1212/01.wnl.0000078928.20107.52

11. Ringholz GM, Appel SH, Bradshaw M, Cooke NA, Mosnik DM, Schulz PE. Prevalence and patterns of cognitive impairment in sporadic ALS. Neurology. 2005;65(4):586–590. doi:10.1212/01.wnl.0000172911.39167.b6

12. Rippon GA, Scarmeas N, Gordon PH, et al. An Observational Study of Cognitive Impairment in Amyotrophic Lateral Sclerosis. Archives of Neurology. 2006;63(3):345–352. doi:10.1001/archneur.63.3.345

13. Montuschi A, Iazzolino B, Calvo A, et al. Cognitive correlates in amyotrophic lateral sclerosis: a population-based study in Italy. J Neurol Neurosurg Psychiatry. 2015;86(2):168–173. doi:10.1136/jnnp-2013-307223

14. Phukan J, Elamin M, Bede P, et al. The syndrome of cognitive impairment in amyotrophic lateral sclerosis: a population-based study. J Neurol Neurosurg Psychiatry. 2012;83(1):102–108. doi:10.1136/jnnp-2011-300188

15. McHutchison CA, Wuu J, McMillan CT, et al. Temporal course of cognitive and behavioural changes in motor neuron diseases. J Neurol Neurosurg Psychiatry. 2024;95(4):316–324. doi:10.1136/jnnp-2023-331697

16. McMillan CT, Wuu J, Rascovsky K, et al. Defining Cognitive Impairment in Amyotrophic Lateral Sclerosis: An Evaluation of Empirical Approaches. Amyotroph Lateral Scler Frontotemporal Degener. 2022;23(7-8):517–526. doi:10.1080/21678421.2022.2039713

17. Johnson JK, Diehl J, Mendez MF, et al. Frontotemporal lobar degeneration: demographic characteristics of 353 patients. Arch Neurol. 2005;62(6):925–930. doi:10.1001/archneur.62.6.925

18. Rosso SM, Donker Kaat L, Baks T, et al. Frontotemporal dementia in The Netherlands: patient characteristics and prevalence estimates from a population-based study. Brain. 2003;126(Pt 9):2016–2022. doi:10.1093/brain/awg204

19. Burrell JR, Kiernan MC, Vucic S, Hodges JR. Motor neuron dysfunction in frontotemporal dementia. Brain. 2011;134(Pt 9):2582–2594. doi:10.1093/brain/awr195

20. Lomen-Hoerth C, Anderson T, Miller B. The overlap of amyotrophic lateral sclerosis and frontotemporal dementia. Neurology. 2002;59(7):1077–1079. doi:10.1212/WNL.59.7.1077

21. Gijselinck I, Van Langenhove T, van der Zee J, et al. A C9orf72 promoter repeat expansion in a Flanders-Belgian cohort with disorders of the frontotemporal lobar degeneration-amyotrophic lateral sclerosis spectrum: a gene identification study. Lancet Neurol. 2012;11(1):54–65. doi:10.1016/S1474-4422(11)70261-7

22. Moore KM, Nicholas J, Grossman M, et al. Age at symptom onset and death and disease duration in genetic frontotemporal dementia: an international retrospective cohort study. Lancet Neurol. 2020;19(2):145–156. doi:10.1016/S1474-4422(19)30394-1

23. Simón-Sánchez J, Dopper EGP, Cohn-Hokke PE, et al. The clinical and pathological phenotype of C9ORF72 hexanucleotide repeat expansions. Brain. 2012;135(3):723–735. doi:10.1093/brain/awr353

24. Samra K, MacDougall AM, Peakman G, et al. Motor symptoms in genetic frontotemporal dementia: developing a new module for clinical rating scales. J Neurol. 2023;270(3):1466–1477. doi:10.1007/s00415-022-11442-y

25. Mahoney CJ, Beck J, Rohrer JD, et al. Frontotemporal dementia with the C9ORF72 hexanucleotide repeat expansion: clinical, neuroanatomical and neuropathological features. Brain. 2012;135(Pt 3):736–750. doi:10.1093/brain/awr361

26. Byrne S, Elamin M, Bede P, et al. Cognitive and clinical characteristics of patients with amyotrophic lateral sclerosis carrying a C9orf72 repeat expansion: a population-based cohort study. Lancet Neurol. 2012;11(3):232–240. doi:10.1016/S1474-4422(12)70014-5

27. Van Langenhove T, Piguet O, Burrell JR, et al. Predicting Development of Amyotrophic Lateral Sclerosis in Frontotemporal Dementia. Journal of Alzheimer’s Disease. 2017;58(1):163–170. doi:10.3233/JAD-161272

28. Smith BN, Newhouse S, Shatunov A, et al. The C9ORF72 expansion mutation is a common cause of ALS+/−FTD in Europe and has a single founder. Eur J Hum Genet. 2013;21(1):102–108. doi:10.1038/ejhg.2012.98

29. Nagao S, Yokota O, Nanba R, et al. Progressive supranuclear palsy presenting as primary lateral sclerosis but lacking parkinsonism, gaze palsy, aphasia, or dementia. J Neurol Sci. 2012;323(1-2):147–153. doi:10.1016/j.jns.2012.09.005

30. Josephs KA, Katsuse O, Beccano-Kelly DA, et al. Atypical Progressive Supranuclear Palsy With Corticospinal Tract Degeneration. Journal of Neuropathology & Experimental Neurology. 2006;65(4):396–405. doi:10.1097/01.jnen.0000218446.38158.61

31. Toledo JB, Van Deerlin VM, Lee EB, et al. A platform for discovery: The University of Pennsylvania Integrated Neurodegenerative Disease Biobank. Alzheimers Dement. 2014;10(4):477–484.e1. doi:10.1016/j.jalz.2013.06.003

32. Xie SX, Baek Y, Grossman M, et al. Building An Integrated Neurodegenerative Disease Database At An Academic Health Center. Alzheimers Dement. 2011;7(4):e84–e93. doi:10.1016/j.jalz.2010.08.233

33. Mackenzie IRA, Neumann M, Baborie A, et al. A harmonized classification system for FTLD-TDP pathology. Acta Neuropathol. 2011;122(1):111–113. doi:10.1007/s00401-011-0845-8

34. Mackenzie IRA, Neumann M, Bigio EH, et al. Nomenclature and nosology for neuropathologic subtypes of frontotemporal lobar degeneration: an update. Acta Neuropathol. 2010;119(1):1–4. doi:10.1007/s00401-009-0612-2

35. Gordon PH, Corcia P, Lacomblez L, et al. Defining Survival as an Outcome Measure in Amyotrophic Lateral Sclerosis. Archives of Neurology. 2009;66(6):758–761. doi:10.1001/archneurol.2009.1

36. Grambsch PM, Therneau TM. Proportional Hazards Tests and Diagnostics Based on Weighted Residuals. Biometrika. 1994;81(3):515–526. doi:10.2307/2337123

37. Snowden JS, Harris J, Richardson A, et al. Frontotemporal dementia with amyotrophic lateral sclerosis: A clinical comparison of patients with and without repeat expansions in C9orf72. Amyotrophic Lateral Sclerosis and Frontotemporal Degeneration. 2013;14(3):172–176. doi:10.3109/21678421.2013.765485

38. Toyoshima Y, Tan CF, Kozakai T, Tanaka M, Takahashi H. Is motor neuron disease-inclusion dementia a forme fruste of amyotrophic lateral sclerosis with dementia? An autopsy case further supporting the disease concept. Neuropathology. 2005;25(3):214–219. doi:10.1111/j.1440-1789.2005.00599.x

39. Hu WT, Seelaar H, Josephs KA, et al. Survival Profiles of Patients With Frontotemporal Dementia and Motor Neuron Disease. Archives of Neurology. 2009;66(11):1359–1364. doi:10.1001/archneurol.2009.253

40. Brettschneider J, Del Tredici K, Irwin DJ, et al. Sequential distribution of pTDP-43 pathology in behavioral variant frontotemporal dementia (bvFTD). Acta Neuropathol. 2014;127(3):423–439. doi:10.1007/s00401-013-1238-y

41. Brettschneider J, Arai K, Del Tredici K, et al. TDP-43 pathology and neuronal loss in amyotrophic lateral sclerosis spinal cord. Acta Neuropathol. 2014;128(3):423–437. doi:10.1007/s00401-014-1299-6

42. Geser F, Prvulovic D, O’Dwyer L, et al. On the development of markers for pathological TDP-43 in amyotrophic lateral sclerosis with and without dementia. Progress in Neurobiology. 2011;95(4):649–662. doi:10.1016/j.pneurobio.2011.08.011

43. Goldman JS, Farmer JM, Van Deerlin VM, Wilhelmsen KC, Miller BL, Grossman M. Frontotemporal dementia: genetics and genetic counseling dilemmas. Neurologist. 2004;10(5):227–234. doi:10.1097/01.nrl.0000138735.48533.26

44. Rohrer JD, Guerreiro R, Vandrovcova J, et al. The heritability and genetics of frontotemporal lobar degeneration. Neurology. 2009;73(18):1451–1456. doi:10.1212/WNL.0b013e3181bf997a

45. Byrne S, Walsh C, Lynch C, et al. Rate of familial amyotrophic lateral sclerosis: a systematic review and meta-analysis. J Neurol Neurosurg Psychiatry. 2011;82(6):623–627. doi:10.1136/jnnp.2010.224501

46. Rhodes E, Alfa S, Jin HA, et al. Cognitive reserve in ALS: the role of occupational skills and requirements. Amyotroph Lateral Scler Frontotemporal Degener. Published online April 9, 2024:1–10. doi:10.1080/21678421.2024.2336113

47. Massimo L, Zee J, Xie SX, et al. Occupational attainment influences survival in autopsy-confirmed frontotemporal degeneration. Neurology. 2015;84(20):2070–2075. doi:10.1212/WNL.0000000000001595

48. Placek K, Baer GM, Elman L, et al. UNC13A polymorphism contributes to frontotemporal disease in sporadic amyotrophic lateral sclerosis. Neurobiol Aging. 2019;73:190–199. doi:10.1016/j.neurobiolaging.2018.09.031

49. Placek K, Benatar M, Wuu J, et al. Machine learning suggests polygenic risk for cognitive dysfunction in amyotrophic lateral sclerosis. EMBO Mol Med. 2021;13(1):e12595. doi:10.15252/emmm.202012595

